# *Bacterial* shedding and serologic responses following an outbreak of *Salmonella* Typhi in an endemic cohort

**DOI:** 10.1101/2021.12.17.21267824

**Authors:** Peter Johnston, Patrick Bogue, Angeziwa Chunga Chirambo, Maurice Mbewe, Reenesh Prakash, Vanessa Kandoole-Kabwere, Rebecca Lester, Thomas Darton, Stephen Baker, Melita Gordon, James Meiring

## Abstract

**Background:** Salmonella enterica serovar Typhi (*S*. Typhi), the causative agent of Typhoid fever, is transmitted faecal-orally. Some typhoid sufferers shed *S*. Typhi beyond convalescence, but culturing stool following every case is impractical. Here we hypothesised that serology might direct testing and identify shedding after a typhoid outbreak.

**Methodology/Principle Findings:** In 2016 there was a typhoid outbreak in a Nursing School in Malosa, Malawi. We collected serum three and six-months post-outbreak. We measured IgG antibody titres against Vi capsular polysaccharide (anti-Vi IgG) and IgM / IgG antibodies against H:d flagellin (anti-H:d). We screened faecal samples from participants with high and low anti-Vi IgG (measured at visit one) by culture and PCR. Participants reported whether they had persistent fever for ≥ three days (in keeping with World Health Organization definitions for typhoid) during the outbreak. We tested for environmental *S*. Typhi.

368 people provided serum at 3-months, of whom 320 provided serum at 6-months; 49 participants provided a faecal sample (25 from the highest and 24 from the lowest deciles for anti-Vi IgG titre). We did not grow *S*. Typhi from faeces, but one sample produced a positive PCR amplification for *S*. Typhi. Median anti-Vi IgG titre fell amongst participants with persistent fever (8.08 to 3.7 EU/ml, *<0.000001, Wilcoxon signed rank*). Median anti-H:d IgG titres fell in those with and without persistent fever (87.8 to 77.4 EU/, *p* = <0.000001 and 82.4 to 79.2 EU/ml, *p = 0.0002, Wilcoxon signed rank, respectively)*. Anti-H:d IgM titres did not change significantly. Non-Typhoidal Salmonellae were identified in water sampled at source and a kitchen tap.

**Conclusions / Significance:** We did not identify culture-confirmed shedding through sero-surveillance. Serologic trends signify a fall from an outbreak-associated peak. Despite effective vaccines, identifying ways to detect and treat shedding remain vital to break transmission and eliminate typhoid.

**Author Summary:** Typhoid fever spreads by the faecal-oral route. Some people continue to shed the bacterium that causes typhoid (*Salmonella* enterica serovar Typhi, or *S*. Typhi) after recovering from the illness. To stop onward spread it is important that these people are identified and treated.

Shedders are detected when *S*. Typhi grows from faeces, but it is not practical to obtain stool samples from large populations. Following a typhoid outbreak we tested whether a subset of participants with high antibodies to *S*. Typhi proteins contained more shedders than a subset with low antibody responses. We tested whether antibody levels changed in the months after the outbreak, to inform whether they are useful markers of exposure in a population.

We did not grow *S*. Typhi. This may be because our population had few risk factors for *S*. Typhi carriage, or because exposure to other endemic bacteria influence antibody levels. We saw a decline in antibody levels over time, most marked in those who reported fever during the outbreak. We think that this reflects a response to recent infection. It is important to continue to evaluate ways of finding carriers so that, combined with vaccines and improved sanitation, we can one day eliminate typhoid.

## Background

It is estimated that 1.5 million cases of typhoid fever occurred in Sub-Saharan Africa in 2017 (1). Whilst this figure represents a fall in incidence estimates from 1990, localised outbreaks (2-5) map closely to the emergence of genotypes associated with multi-drug resistance (MDR) (6). Mathematical modelling suggests such outbreaks may be driven by increased transmissibility associated with drug resistance phenotypes (7). Where *Salmonella* enterica serovar Typhi (*S*. Typhi) is inadequately treated, ongoing shedding becomes more likely, and detection of this is essential in order to break the transmission cycle.

*S*. Typhi is shed intermittently in stool, and bacterial culture lacks sensitivity (8). A recent case-control study of children living in Nairobi found *S*. Typhi carriage rates of 1.1% among asymptomatic individuals (9). The relative contribution of temporary shedders to ongoing transmission remains unclear, but is likely to play a larger role in high incidence settings (10, 11).

In non-endemic settings, serology has been exploited to complement an outbreak investigation, with candidate shedders successfully identified by a high anti-Vi antibody titre (anti-Vi IgG) followed by isolation of *S*. Typhi in stool (12, 13). Serology has been used in attempts to identify carriers in endemic populations, but has shown limited success (14-16).

In June 2016 an outbreak of febrile illness occurred in Malosa, Malawi. Three patients presenting to the local hospital had blood culture-confirmed typhoid fever. Cases were traced to a local primary school, secondary school and a residential nursing school. Over the next month the District Health Office identified 101/407 nursing school residents who met a clinical case definition for typhoid fever.

Our primary objective was to identify nursing school residents shedding *S*. Typhi after the outbreak, using serology to target subsequent testing of high risk individuals. We focussed our efforts on the nursing school because students continuing to shed *S*. Typhi might transmit typhoid to patients. Secondary objectives were to identify any environmental source of *S*. Typhi within the nursing school and to explore serologic responses to typhoid exposure, as determined by self-reported, persistent fever.

## Methods

We performed a prospective observational study comprising two study visits, conducted three and six months after the Malosa outbreak.

Any person over 18 years of age who had been resident in the nursing school between 1^st^ June 2016 and 31^st^ August 2016 was eligible to participate. We enrolled participants and conducted study visit one simultaneously in October 2016, three months after the outbreak.

We collected a blood sample from each participant and completed a structured case record. This included self-report of whether the participant had a persistent fever (defined as three days or more duration) during the outbreak. Those with persistent fever were considered typhoid cases based on the WHO case finding definition (17).

We identified participants whose three-month anti-Vi IgG levels were in the top ten percent of the cohort and participants whose anti-Vi IgG were in the lowest ten percent of the cohort. These participants were approached and asked to provide a stool sample. Samples were collected between the three- and six-month study visits, after anti-Vi IgG results became available. Stool was sent for culture and polymerase chain reaction (PCR) testing.

The entire cohort was approached again in January 2017, six months after the outbreak. A further blood sample was obtained for repeat serology.

### Laboratory procedures

We determined anti-Vi IgG antibody concentration using a commercial Enzyme-linked immunosorbent assay (ELISA) in accordance with manufacturer’s instructions (VaccZyme™ The Binding Site, Birmingham, UK). We derived antibody concentration in ELISA units/ml (EU/ml) from the optical density using a standardised curve-fitting logistic method.

Anti-H:d IgM and IgG were performed at the Oxford University Clinical Research Unit in Vietnam, using in-house ELISAs, as previously described (18).

Stool samples were collected in sterile sample pots and transferred to the laboratory on the same day. Stool was pre-enriched in Selenite broth before being sub-cultured on Xylose Lysine Deoxyxholate (XLD) agar at 37°C for 18-24 hours. Single colonies from XLD were sub-cultured on MacConkey and Sheep blood agar (SBA) plates. Colonies from SBA plates were used for biochemical tests. Analytical Profile Index (API 20E) tests were used for *Salmonella* identification. Following the White-Kauffmann-Le Minor scheme, serotyping was done using polyvalent O and H, O4, O9, Hd, Hg, Hi, Hm, and Vi antisera (Pro-Lab Diagnostics).

PCR testing was used alongside routine culture to increase the yield from stool. DNA was extracted from Selenite broth pre-enriched stool specimens using QIAamp Fast DNA Stool Mini Kit (Qiagen). A pan *Salmonella* PCR targeting the *ttr* gene, encoding tetrathionate reductase, and a multiplex PCR with a pan-*Salmonella* invasion A gene, *S*. Typhi fimbriae gene, and the kit’s internal control were performed (19, 20).

### Environmental sampling

Malosa Dam provides water for the nursing school and secondary school via a gravity fed water pipe feeding in to a holding tank. The water is reportedly chlorinated daily after leaving the holding tank.

We took 36 environmental samples, in-duplicate, over two days. These comprised stool from food-handlers, food and tap water from the kitchens and water from the dam. Samples were cultured and sero-typed as described above.

### Statistical methods

We treated serologic data as a metric continuous variable, and our surrogate measure of typhoid exposure as dichotomous (fever for ≥ 3 days or not).

We tested continuous variables for normality using the Shapiro-Wilks test before and after log-transformation. We used nonparametric tests where data were not normally distributed.

We used paired significance tests to compare change in antibody titre within the population as a whole and change in antibody titre depending upon whether the clinical definition for typhoid had been met. We used unpaired significance tests to compare antibody titres between participants who did and did not meet the WHO definition for typhoid at each study visit. Only participants who submitted serum at three and six months contributed to paired analyses.

Statistical analyses were carried out using the free to use, open source statistical package ‘R’ version 4.1.1 (21). The significance threshold was set at 0.05 for all tests.

### Ethical considerations

All participants provided written informed consent. Participant information materials were made available in English and Chichewa languages. Our protocol stated that those who were stool culture positive for *S*. Typhi would be advised by the District Health Office to take either 28 days of ciprofloxacin or 14 days of azithromycin with the aim of eradicating carriage and preventing onward transmission. This study was approved by College of Medicine Research Ethics Committee, P.10/16/2043.

## Results

We recruited 374 participants from a total population of 407. 368 participants submitted blood samples three months post outbreak. 320 participants submitted blood at three months and six months. We obtained stool samples from 25 of 36 participants whose anti-Vi IgG response was in the highest decile, and from 24 of 36 participants whose anti-Vi IgG response was in the lowest decile. The characteristics of included participants are shown in Table one.

**Table 1.** Baseline characteristics of participants who submitted blood samples at study visit one

|  | Clinical typhoid | No clinical typhoid |  |
| --- | --- | --- | --- |
| Participants (N) | 156 | 212 |  |
| Age in years (mean) | 22.6 | 24.2 |  |
| (range) | 8-54 | 18-42 |  |
| (SD) | 4.4 | 6.3 |  |
| gender (proportion female, (%)) | 78/156 (50) | 106/212 (50) |  |
| Treated with antibiotics (%) | 140/156 (90) | 16/212 (7.5) |  |
| Median anti-Vi IgG titre at visit one (EU/ml) | 8.08 (IQR 17.9) | 3.7 (IQR 12.4) | *P* =0.11 (Mann-Whitney-U test) |
| Median anti-H:d IgM antibody titre at visit one (EU/ml) | 55.2 (IQR 51.4) | 52.1 (IQR 55.9) | *P*=0.34 (Mann-Whitney-U test) |
| Median anti-H:d IgG antibody titre at visit one (EU/ml) | 87.8 (IQR 85.1) | 82.4 (IQR 88.4) | *P* = 0.27 (Mann-Whitney-U test) |
| Median anti-Vi IgG titre at visit two (EU/ml) | 3.7 (IQR 17.7) | 3.7 (IQR 14.4) | *P*=0.86 (Mann-Whitney-U test) |
| Median anti-H:d IgM antibody titre at visit two (EU/ml) | 54.2 (IQR 44.9) | 55.3 (IQR 56.1) | *P*=0.92 (Mann-Whitney-U test) |
| Median anti-H:d IgG antibody titre at visit two (EU/ml) | 77.4 (IQR 57) | 79.2 (IQR 65.7) | *P*=0.80 (Mann-Whitney-U test) |
| Headache, N (%) | 139/156 (89) | 39/212 (18.4) |  |
| Abdominal pain, N (%) | 82/156 (52.6) | 25/212 (11.8) |  |
| Diarrhoea, N (%) | 62/156 (39.7) | 29/212 (13.7) |  |
| Constipation, N (%) | 31/156 (19.9) | 10/212 (4.7) |  |
| Nausea, N (%) | 60/156 (38.5) | 13/212 (6.1) |  |

### Stool Culture

Of 49 participants who submitted stool for culture, none were culture positive for *S*. Typhi in stool. Four participants had Salmonella spp. in stool on routine culture. Three of these participants were in the lowest decile for anti-Vi IgG level at visit one, one was in the highest decile. None of the stool samples grew *S*. Typhi (Figure 1). *Ttr* and multiplex PCR test did not detect any additional *Salmonella* from the stool samples. One Salmonella isolate was identified as *S*. Typhi using the *S*. Typhi fimbriae gene target on the multiplex PCR. This participant had the highest three-month anti-Vi IgG and anti-H:d IgG titres among participants submitting stool. (Supplemental Figures 1-3).

**Figure 1.**
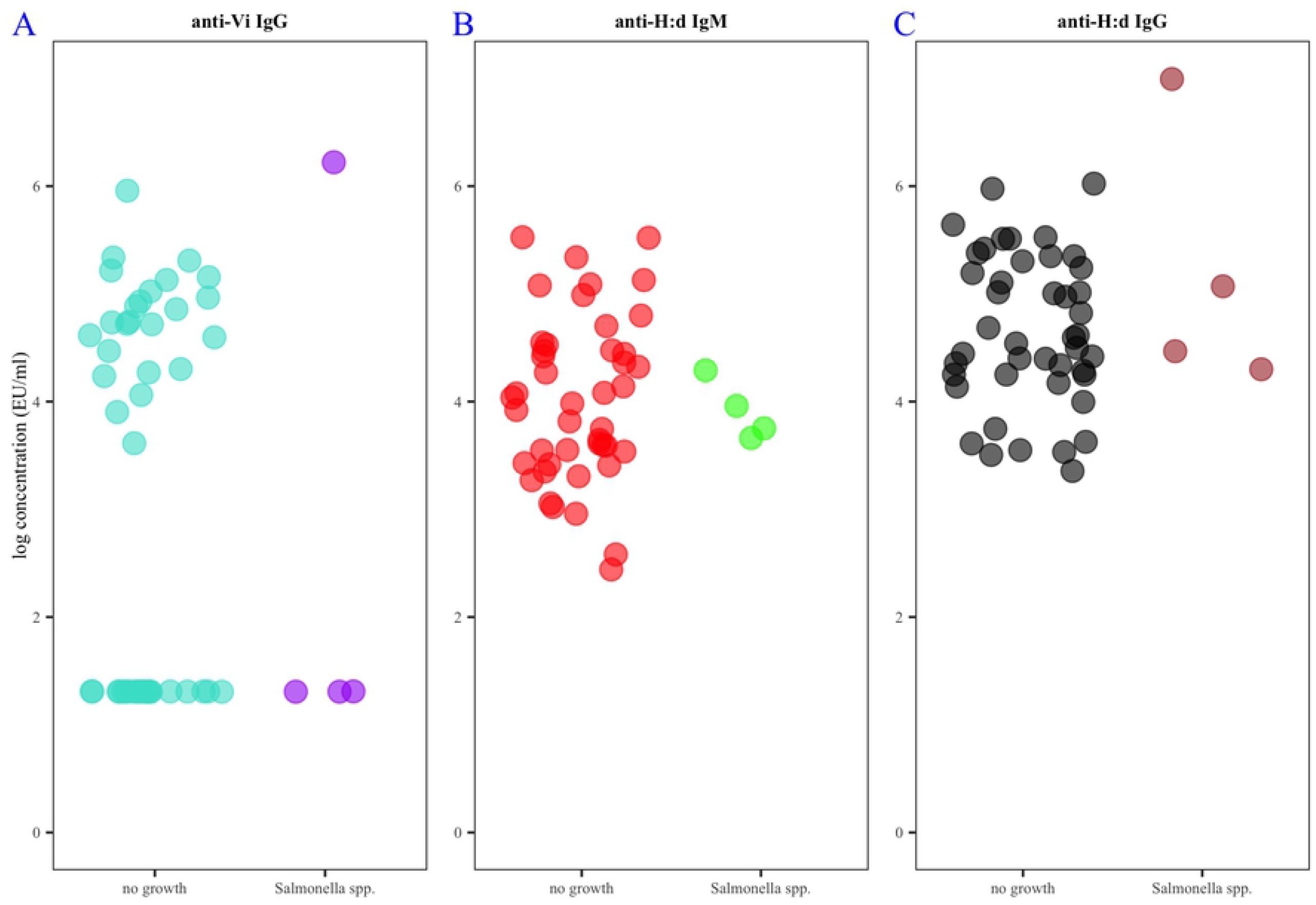
**A**. Log anti-Vi IgG concentration at the three month visit among participants who grew Salmonella spp. in stool (purple) compared with those who had no growth (turquoise). **B**. log anti-H:d IgM concentration at the three month visit among participants who grew Salmonella spp. in stool (green) compared with those who did not (red). **C**. log anti-H:d IgG concentration at the three month visit among participants who grew Salmonella spp. in stool (brown) and those who did not (black).

### Antibody responses

Among participants with persistent fever there was a significant fall in anti-Vi IgG between three and six months (median titre 8.08 to 3.7 EU/ml (*p = <0*.*000001, Wilcoxon signed rank test*). This was not true of participants without persistent fever (median titre 3.7 to 3.7 EU/ml, *p = 0*.*12, Wilcoxon signed rank test*) (Figure 2.)

**Figure 2.**
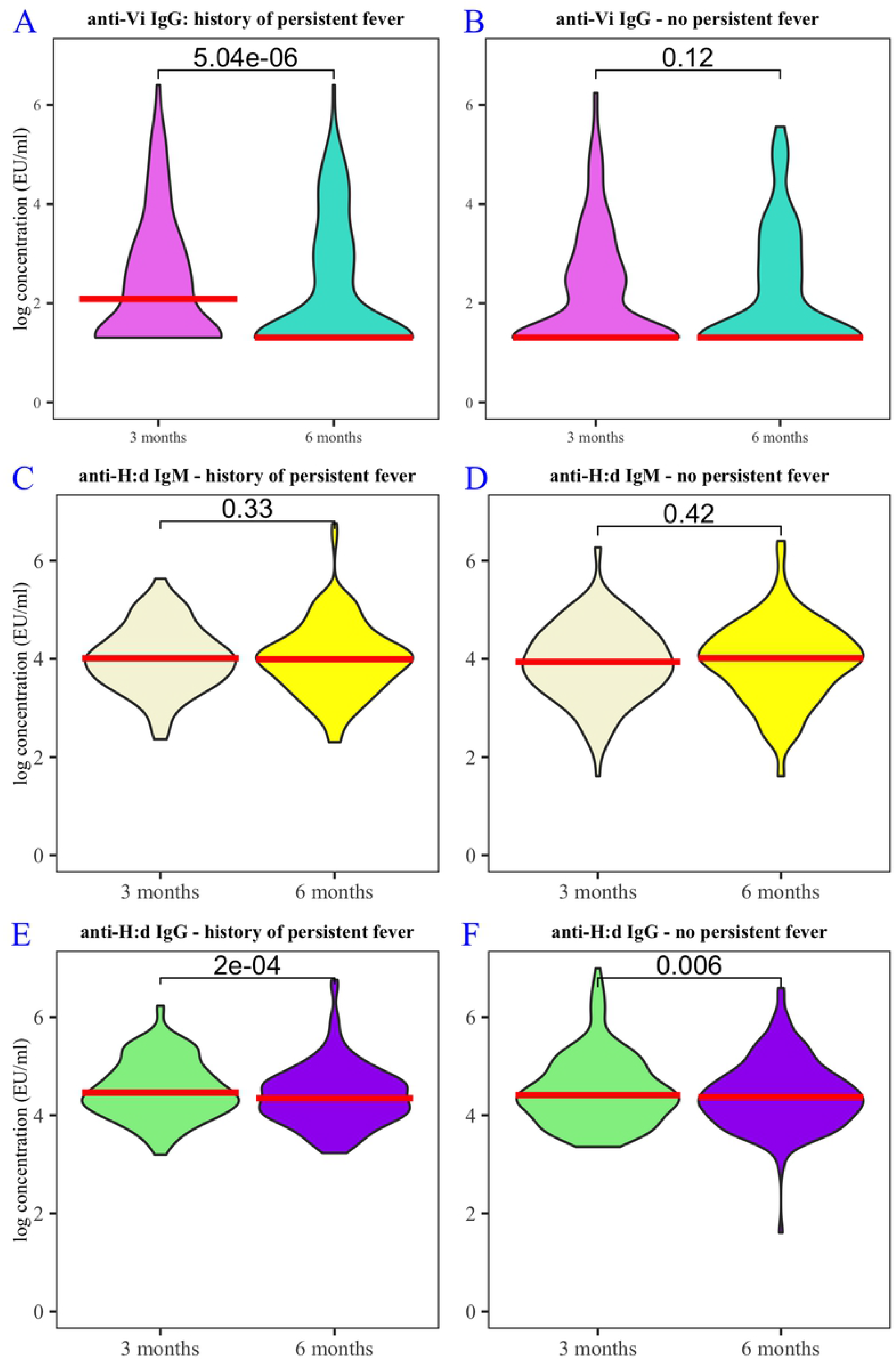
**A**. log anti-Vi IgG concentration amongst participants with persistent fever, comparing titres at 3 months (lilac) and 6 months (turquoise) post-outbreak. **B**. log anti-Vi IgG concentration amongst participants without persistent fever, comparing titres at 3 months (lilac) and 6 months (turquoise) post-outbreak. **C**. log anti-H:d IgM concentration amongst participants with persistent fever, comparing titres at 3 months (beige) and 6 months (yellow) post-outbreak. **D**. log anti-H:d IgM concentration amongst participants without persistent fever, comparing titres at 3 months (beige) and 6 months (yellow) post outbreak. **E**. log anti-H:d IgG concentration amongst participants without persistent fever, comparing titres at 3 months (green) and 6 months (purple) post-outbreak. **F**. log anti-H:d IgM concentration amongst participants without persistent fever, comparing titres at 3 months (green) and 6 months (purple) post-outbreak. All graphs: red crossbar shows median antibody concentration; brackets show p-values (Wilcoxon signed rank test).

Among participants with persistent fever there was a significant fall in anti-H:d IgG between three and six months (median titre 87.8 to 77.4 EU/ml (*p = 0*.*0002, Wilcoxon signed rank test*). Among participants without persistent fever, there was also a significant fall in anti-H:d IgG between three and six months (median titre 82.4 to 79.2 EU/ml, *p = 0*.*006, Wilcoxon signed rank test*) (Figure 2.)

There was no significant change in anti-H:d IgM between three and six months, either among those with persistent fever (median titre 55.2 to 54.2 EU/ml (*p = 0*.*33, Wilcoxon signed rank test*) or without persistent fever (median titre 52.1 to 55.3 EU/ml (*p = 0*.*42, Wilcoxon signed rank test*). (Figure 2.)

### Environmental Sampling

Non-typhoidal *Salmonella* was identified from tap water in the kitchen of the nursing school and in water from Malosa dam. No *S*. Typhi was identified.

## Discussion

Our study shows that, even in a population enriched through the circumstances of an outbreak, sero-surveillance could not identify culture-confirmed shedding of *S*. Typhi. Sero-surveillance combined with PCR may have identified one participant shedding *S*. Typhi. Anti-Vi IgG titres fell amongst participants reporting persistent fever during the outbreak. Anti-H:d IgG titres fell amongst participants with and without persistent fever. There was no change in anti-H:d IgM, irrespective of fever status. We did not find an environmental reservoir of *S*. Typhi, but non-typhoidal *salmonellae* were present in water samples from the reservoir supplying the nursing school, and a tap in the kitchen. The presence of these bacteria indicate contamination and under-treatment of the water supply, and represent a possible source of the outbreak.

As our population was pre-defined, we could not perform an *a priori* power calculation. Our study population was young (mean 23.5 years). We expect low rates of biliary pathology in the population based on discussions with surgical colleagues working in the area. A similar study design might have detected *S*. Typhi shedding amongst an older cohort, or in an endemic setting where rates of biliary disease are high.

Participants from the Nursing College were on clinical placement in multiple institutions across Southern Malawi. This made enrolment and follow-up challenging: we were unable to obtain stool samples from all the participants that we identified in the highest and lowest anti-Vi IgG titre groups. Given the intermittent nature of shedding, several samples over time would have increased our chances of detecting *S*. Typhi by culture. One participant was identified as having PCR amplification of the *fimbriae* target. This suggests that they may have been carrying / shedding *S*. Typhi, but this could not be verified through culture confirmation. This participant had the highest anti-Vi IgG and anti-H:d IgG titres of all those who submitted stool.

The fall in anti-Vi IgG and anti-H:d IgG suggests that a peak titre is reached within the post outbreak surveillance period encompassed by our study, and that titres have fallen six months post exposure. It is possible that anti-Vi IgG is more specific to febrile typhoid illness, or that anti-H:d IgG falls more swiftly after exposure and so an effect was demonstrable across the entire cohort. This would be in keeping with findings from a human challenge model (22). The static anti-H:d IgM concentration suggests that any change in titre precipitated by the outbreak has been and gone at three months.

The Vi (virulence) antigen comprises a polysaccharide expressed on the surface of *S*. Typhi, as well as *Salmonella* Paratyphi C, Citrobacter *freundii* and *Salmonella* Dublin (23, 24). IgG antibodies to the Vi antigen have been detected in high titres among microbiologically confirmed shedders of *S*. Typhi (14, 25, 26). *S*. Typhi shedders have been identified through their high anti-Vi titre in the context of two outbreaks in the United States (12, 13). Yet the role of anti-Vi IgG for population screening for Typhoid shedding remains unclear: a 2004 study obtained sera from 3209 adults in a Typhoid-endemic area (Vietnam). Multiple rectal swabs were taken from 103 participants with the highest anti-Vi IgG titres at different time points and no *S*. Typhi was isolated (15). Although we obtained fewer faecal specimens, our study population was ‘enriched’ by a known recent exposure to typhoid. The paucity of *S*. Typhi detection in both settings suggests that anti-Vi IgG might be elevated for reasons other than biliary carriage, such as cross-reactivity with other bacteria that express the Vi antigen, or convalescence from acute typhoid fever. It is plausible that both factors would be more significant in areas of the world where typhoid fever is endemic, explaining the geographical discrepancy. A Chilean study comparing anti-Vi IgG amongst bacteriologically proven *S*. Typhi carriers and healthy control subjects and those with acute typhoid fever supports our finding that anti-Vi IgG is a non-specific marker in an endemic setting: although the authors demonstrated higher antibody titre among the shedders, one quarter of chronic carriers did not have a high anti-Vi IgG titre (14).

A recent study utilising culture confirmed carriers who underwent cholecystectomy in Nepal found 13 putative antigens in sera that were expressed highly in the carrier state, and which might bear closer scrutiny in future studies looking for shedding (27). Future studies should focus on robust microbiological confirmation of *S*. Typhi shedding status to provide clear comparators for prospective serologic markers. The detection of carriers will require repeated stool sampling over time, and newer PCR techniques may increase sensitivity over culture alone (19, 28).

Despite the success of conjugate typhoid vaccines in preventing typhoid cases, the recent cluster randomised trial in Bangladesh has yet to show significant indirect protection (29). Wider public health measures, including identifying those who continue to shed *S*. Typhi after infection, remain key to reducing transmission and to ultimately eradicating typhoid.

## Data Availability

All data produced in the present study are available upon reasonable request to the authors

## Supplemental Figures

**Supplemental Figure 1.**
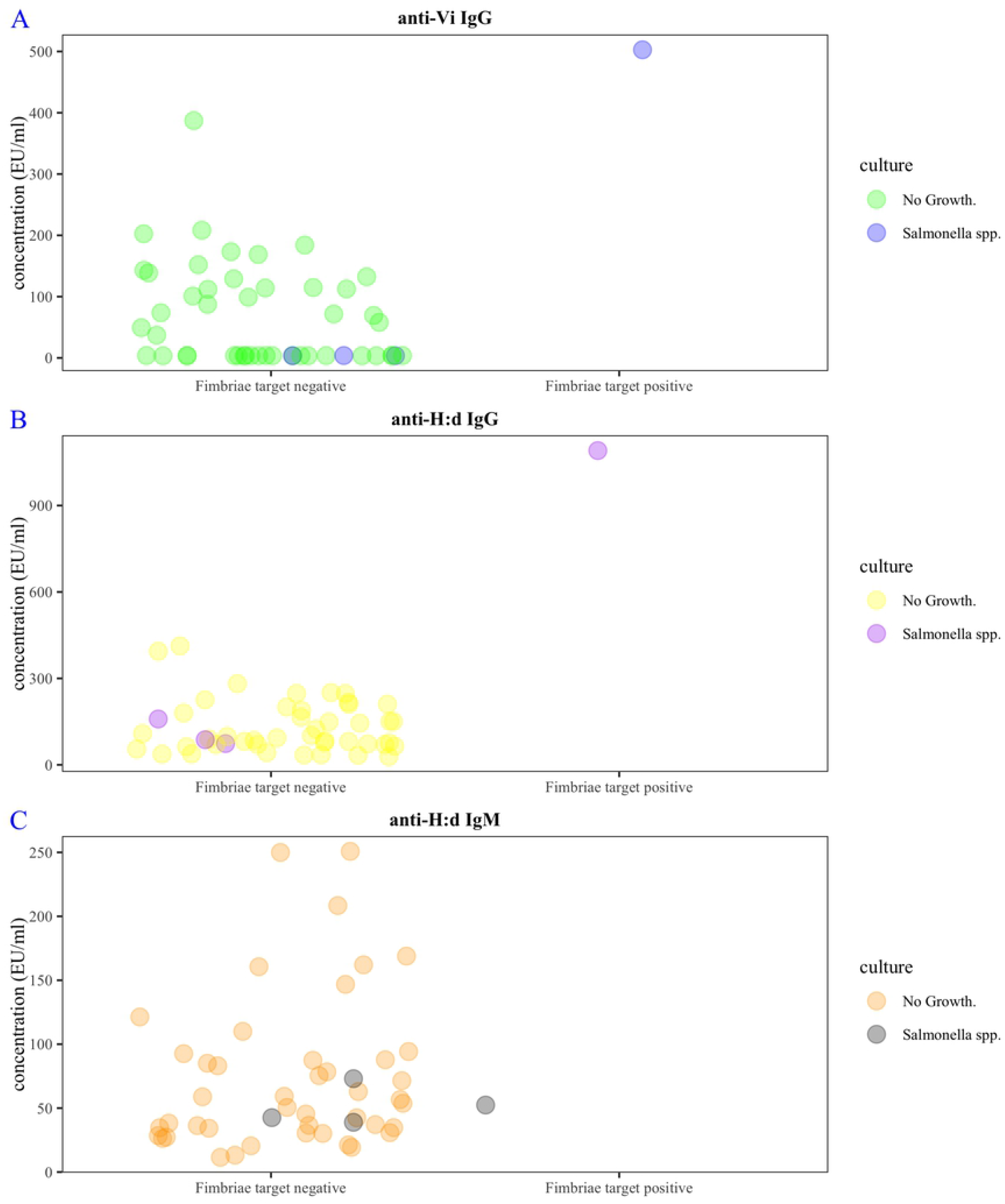
**A**. Log anti-Vi IgG concentration at the three month visit among participants who did and did not amplify the fimbriae PCR target from stool. Green coloured dots represent participants who were culture-negative for Salmonella spp. from stool, turquoise coloured dots represent those who were culture-positive for Salmonella spp from stool. **B**. log anti-H:d IgM at the three month visit among participants who did and did not amplify the fimbriae PCR target from stool. Yellow coloured dots represent participants who were culture-negative for Salmonella spp. from stool, purple coloured dots represent those who were culture-positive for Salmonella spp from stool. **C**. log anti-H:d IgG at the three month visit among participants who did and did not amplify the fimbriae PCR target from stool. Orange coloured dots represent participants who were culture-negative for Salmonella spp. from stool, grey coloured dots represent those who were culture-positive for Salmonella spp from stool.

